# The effects of containment measures in the Italian outbreak of COVID-19

**DOI:** 10.1101/2020.03.25.20042713

**Authors:** M. Supino, A. d’Onofrio, F. Luongo, G. Occhipinti, A. Dal Co

## Abstract

**Background:** The COVID-19 pandemic is spreading worldwide. Italy emerged early on as the country with the largest outbreak outside Asia. The outbreak in Northern Italy demonstrates that it is fundamental to contain the virus’ spread at a very early stage of diffusion. At later stages, no containment measure, even if strict, can prevent the saturation of the hospitals and of the intensive care units in any country.

**Methods and Results:** Here we show that it is possible to predict when the intensive care units will saturate, within a few days from the beginning of the exponential growth of COVID-19 intensive care patients. Using early counts of intensive care patients, we predict the saturation for Lombardy, Italy. We also assess short-term and long-term lockdown effects on intensive care units and number of deaths.

**Conclusions:** Governments should use the Italian outbreak as a precedent and implement appropriate containment measures to prevent the saturation of their intensive care units and protect their population, also, and above all, in anticipation of a possible second exponential spread of infections.

## Introduction

The Coronavirus Disease 2019 (COVID-19) is a respiratory infectious disease caused by the virus SARS-CoV-2 (also known as 2019-nCoV), which originated in Wuhan, China, probably in early December 2019. On January 23^rd^, Wuhan city shut down public transportation and airways; one week later, Wuhan and other cities in the province of Hubei, imposed strict social distancing measures (closure of school and non-essential work activities), combined with active search and isolation of infective cases and their contacts; on February 13^th^, all non-essential companies and manufacturing plants were closed. On February 15^th^, France reported the first death from COVID-19 outside Asia, while dozens of countries document cases of infection. On February 24^th^, three weeks after the lockdown of Wuhan and other cities, cases in China have fallen from an average of 2,500 daily to 400 cases. In the meanwhile, Italy emerged as the country with the largest outbreak outside Asia. On March 9^th^, Italy imposed a lockdown of the whole nation. On March 11^th^, the World Health Organization (WHO) declared the pandemic state, when the world counted more than 118,000 cases in 114 countries. On March 19^th^, while China announced that the incidence was brought to negligible levels and attempts to prudently restart normal life were initiated, the total number of deaths outside China overtook the deaths counted in China. On March 19^th^, Italy counted 3,405 deaths, surpassing China. In Europe, several countries experienced similar exponential growth of cases as Italy, with just a few days of delay (Fig. 1). Spain and France imposed a lockdown on March 14^th^ and on March 17^th^, respectively. On March 21^st^, Italy imposed a full lockdown of the nation, closing all non-essential companies and manufacturing plants. Over the following two weeks, almost all European countries imposed similar containment measures, and more than half of the world’s population was under lockdown, which impacted tremendously societies and economies [1, 2].

**Figure 1.**
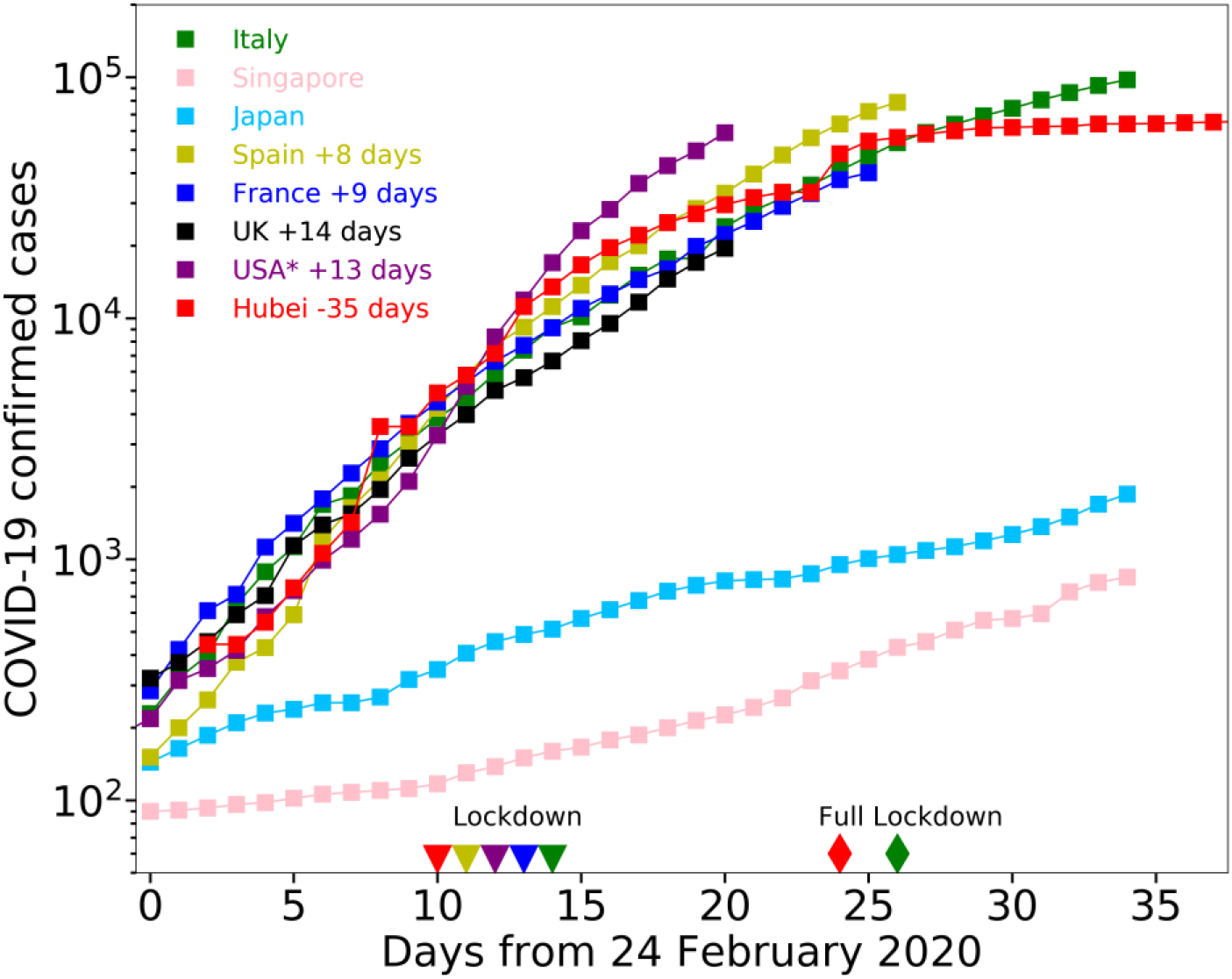
Number of confirmed COVID-19 cases for different countries. Trajectories are shifted temporally to superpose one on another, so that day zero represents the onset of COVID-19 outbreak for all countries. Japan and Singapore were able to contain the outbreak at the very early stage. The Chinese region of Hubei was able to contain diffusion at a later stage, by imposing first a milder lockdown of the population on January 30^th^ 2020 (red triangle), and then a full lockdown of the population on February 13^th^ 2020 (red diamond), closing all non-essential companies and manufacturing plants. Italy, Spain, France, UK, and the American States of California and New York (referred as US*) display exponential growth of confirmed cases. Italy, Spain, France and US* imposed the lockdown at similar (relative) times, indicated by the green, gold, blue and purple triangles, respectively. The four countries have comparable population sizes to Hubei region (58.5M people), with the minimum in Spain (46.7M people) and the maximum in France (66.9M). Italy additionally imposed the full lockdown 12 days afterwards (green diamond).

China was able to control the outbreak of COVID-19 in about two months by implementing strong containments measures, such as lockdown of the population [3]. Lockdown can appear as an extreme measure, but it is not. Mild restrictions, such closure of schools, partial closure of workplaces or curfews, can lower the effective basic reproduction number of infection (R_t_) but very likely not below the unit. With mild restrictions, the number of infections would grow at the same rate for a period equal to the incubation time (which is below 14 days for 99% of COVID-19 infections [4]), and would then grow exponentially at a lower rate. Therefore, mild restrictions would slow down the epidemic, but not control it [5].

In this work, we discuss the necessity and efficacy of lockdown measures for controlling the outbreak of COVID-19, analyzing data from Italy, the country with the first recorded outbreak of the disease during the first spread of COVID-19 in Europe. We also assess the time it takes to reduce the number of patients hospitalized with severe COVID-19 symptoms after the beginning of the lockdown. Currently (October 28^th^, 2020), COVID-19 cases are surging worldwide, following the relaxation of containment measures during the last months. The impact of lockdown in the first Italian outbreak represents a fundamental precedent that can help policymakers to define the decision timeline for containment measures in the next weeks or months.

### COVID-19 represent a severe challenge for National Health Systems

SARS-CoV-2 poses a major challenge for the National Health Systems (NHS): the exponential increase of patients needing hospitalization in intensive care units (ICU). In Italy, the 2018-2019 seasonal influenza required 812 intensive care beds [6], while COVID-19 infection required more than 1000 ICU beds after only 21 days from the first detected cases on February 24^th^ 2020 (Fig. 2).

**Figure 2.**
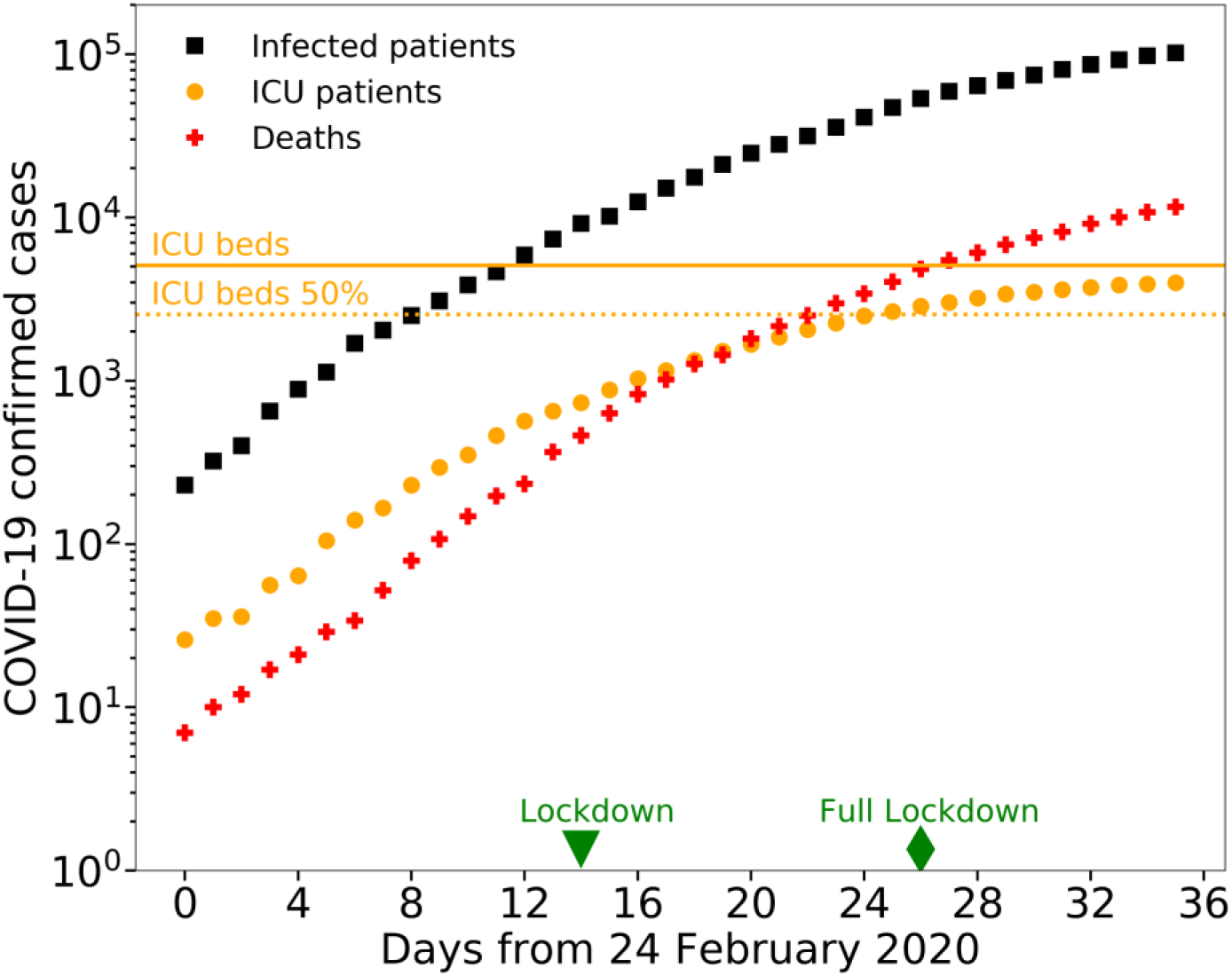
Time evolution of COVID-19 epidemic in Italy. The number of infected patients (black squares) and deaths (red crosses) increased exponentially in time (note that the vertical axis has exponential units). The number of intensive care patients (orange circles) grew exponentially until it reached the maximum capacity of the NHS, and then slowed down as the ICU beds saturated in the Italian regions of Lombardy, Piedmont, Marche, Trentino Alto Adige, Valle d’Aosta, which host 28% of the Italian population. The green triangle and diamond represent the lockdown day (March 9^th^ 2020) and full lockdown day (March 21^st^ 2020) respectively. The orange solid line and dotted line represent 100% and 50% of the total number of ICU beds in Italy before the onset of the epidemic, which was equal to 5090 [7].

While NHS are prepared to receive ICU patients distributed during the influenza season, which lasts several months, no NHS can manage the exponential growing of the number of COVID-19 patients. To avoid the saturation of the ICUs, governments need to impose strong containment measures, such as lockdown of the population early. Acting early is paramount: after the containments measures are taken, the number of cases still grows exponentially for at least ten days, due to infections contracted before the measures [4]. The later these containment measures are taken, the stronger these measures need to be to contain the diffusion of SARS-CoV-2, and could be anyways insufficient to avoid the catastrophic collapse of the NHS. For example, Japan and Singapore were able to avoid the lockdown of the population, because the governments implemented effective measures at the very early stage of the outbreak (Fig. 1).

### COVID-19 epidemic in Italy can be used as a precedent for other countries

From the early stage of the COVID-19 outbreak (February 24^th^ 2020), Italy is providing statistics of the epidemic, through a daily bulletin and an open-access repository [8]. This repository contains daily counts of confirmed cases, hospitalized patients, ICU patients, and deceased patients, at the national and regional level (Fig. 3). This repository represents an important and unique source of information for other countries stroke by the pandemic. The number of ICU patients represents a more robust information compared to the number of infected people, which is subject to under-reporting. The number of infected people strongly depends on the number of performed tests and on the strategy of sampling of the population (e.g. only symptomatic people, randomly chosen people). Testing capability and strategy might vary among different countries largely, while ICU patients count is a routine operation performed by all NHSs. ICU counts are more reliable also compared to deaths counts, since most patients dying with COVID-19 have comorbidities and ascertaining that COVID-19 was the primary cause of death can be complicated. Generally, ICU patients counts offers a far more reliable information of the evolution of COVID-19 epidemic, at least until the ICU capability of the NHS is saturated.

**Figure 3.**
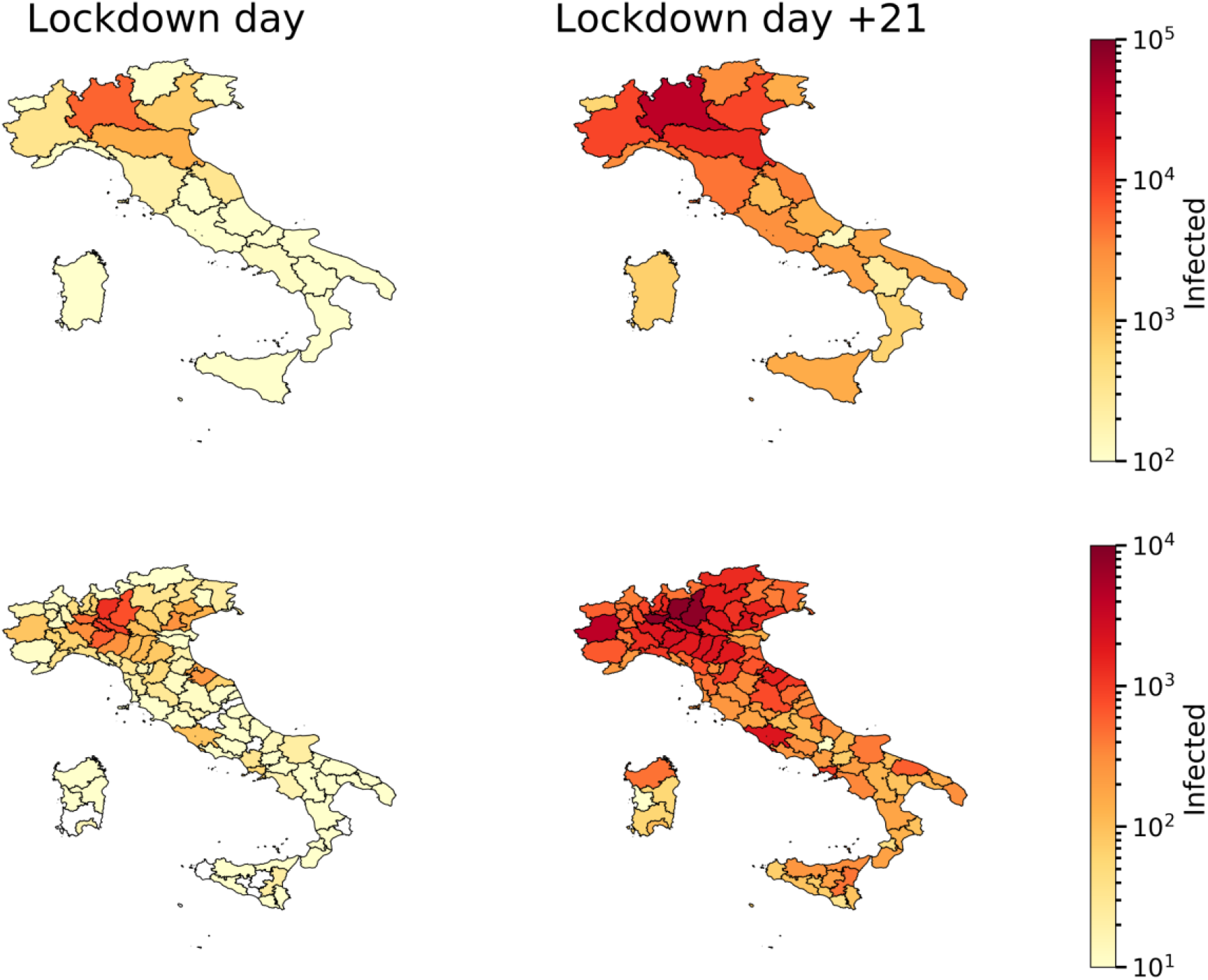
Spatial distribution of COVID-19 epidemic in Italy. The maps on the left represent the number of infection cases detected at the regional level (top panel) and the provincial level (bottom panel), on March 9^th^, when lockdown was imposed. The maps on the right represent the number of total confirmed cases twenty-one days later, on March 30^th^.

### COVID-19 cases evolution in Italy

COVID-19 outbreak in Italy started in Codogno, a town of 15,868 inhabitants in Lombardy region. It is still unclear what events triggered the large outbreak in this region [9, 10]. After ten days from the first ICU patient (February, 20^th^ 2020), the number of COVID-19 ICU patients was already larger than 100; after 18 days from the first ICU patient, ICU patients occupied about the 50% of the total ICU beds of Lombardy region. This percentage is close to the average percentage of ICU beds that are available during the whole year in Italy [7]. At the early stage of the COVID-19 outbreak, the number of ICU beds in Italy was approximately 5090 (i.e. 8,42 ICU beds per 100.000 inhabitants [7]) and Lombardy region had approximately 900 ICU beds (9 ICU beds per 100.000 inhabitants). The Italian government immediately implemented several measures to increase the number of available ICU beds, for example postponing non-essential surgical operations and purchasing mechanical ventilators for new ICUs, and to increase medical support, for example by recalling retired doctors. Yet, these measures offer what we call a *linear response*, which is not sufficient to face an exponential growth of ICU beds demand.

The Italian government implemented also a series of measures to slow down the diffusion of the virus. Schools and universities were closed in Lombardy on February 26^th^, and in rest of Italy on March 5^th^. On March 8^th^, Lombardy region ordered the lockdown of the populations (Decreto del Presidente del Consiglio dei Ministri 8 Marzo 2020). On the following day, the lockdown was extended to the entire nation. In Lombardy and in Italy, nurses, doctors and health-care professionals have fought with incredible strength, acting as war heroes in dark times. However, the ICUs rapidly saturated by COVID-19 ICU patients in Lombardy region (Fig. 4) and subsequently in other regions (Fig. 5). On March 21^st^, the Italian government imposed the full lockdown of the nation, closing off all the non-essential companies and manufacturing plants.

**Figure 4.**
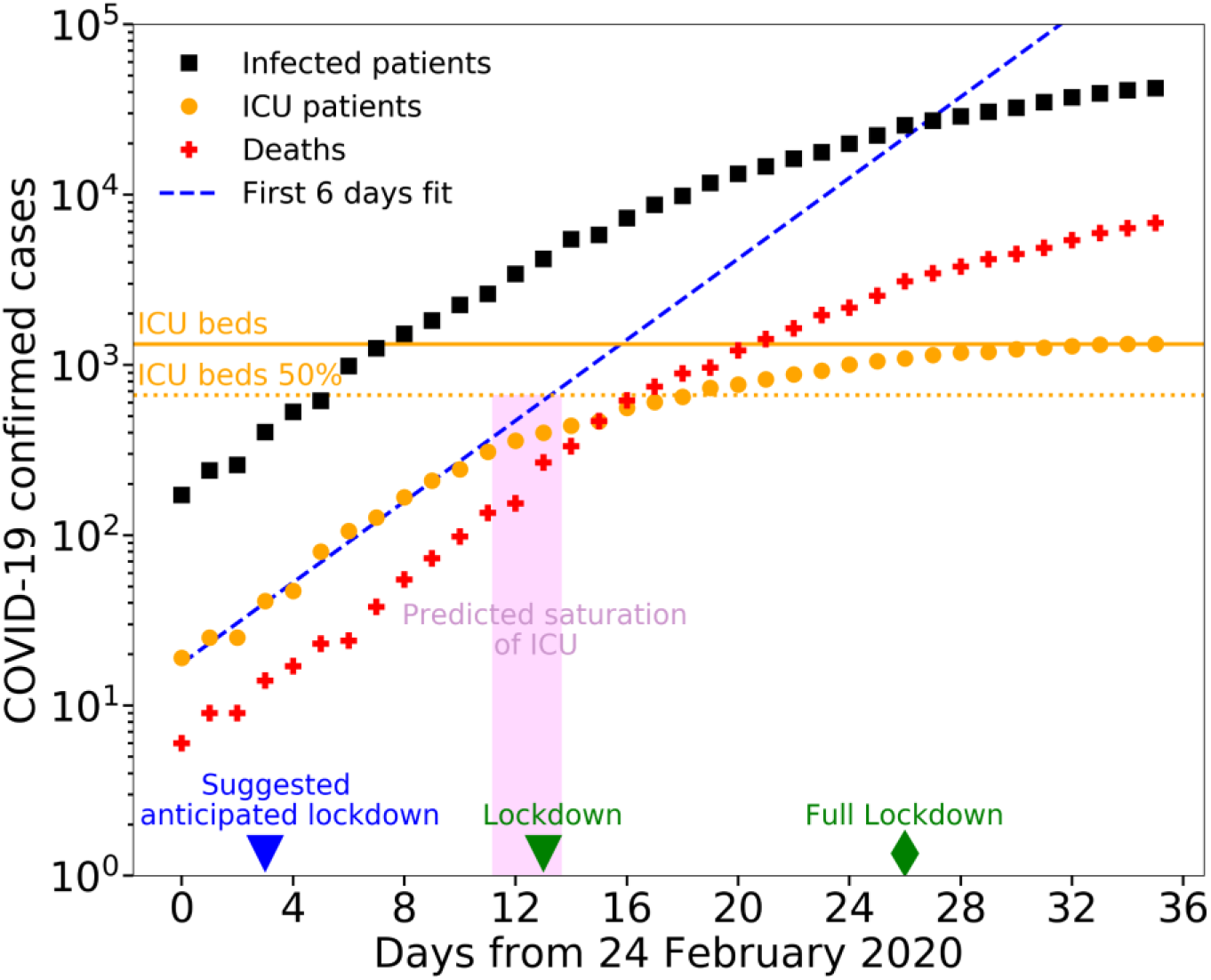
The date of ICUs saturation can be predicted early on. The number of deaths (red crosses) grows exponentially in time in Lombardy region, Italy. The number of ICU patients (orange circles) flattens in time: this indicates the saturation of the ICU beds in the region. The dotted blue line is the linear regression of the first six orange data points. The day of the saturation can be predicted by linear regression, fitting the logarithm of the number of ICU patients at early stages of the epidemic: the timeline highlighted in magenta shows the predicted temporal range for the saturation of 50% of the ICU beds; the beginning and the end of the magenta range are obtained by fitting the first four (blue triangle) and nine orange data points respectively. To avoid ICUs saturation, an earlier lockdown (indicated by the blue triangle) would have been necessary. The green triangle and green diamond represent the lockdown day (March 8^th^ 2020) and full lockdown day (March 21^st^ 2020). The orange solid line and dotted line represent 100% and 50% of the actual total number of ICU beds in Lombardy.

**Figure 5.**
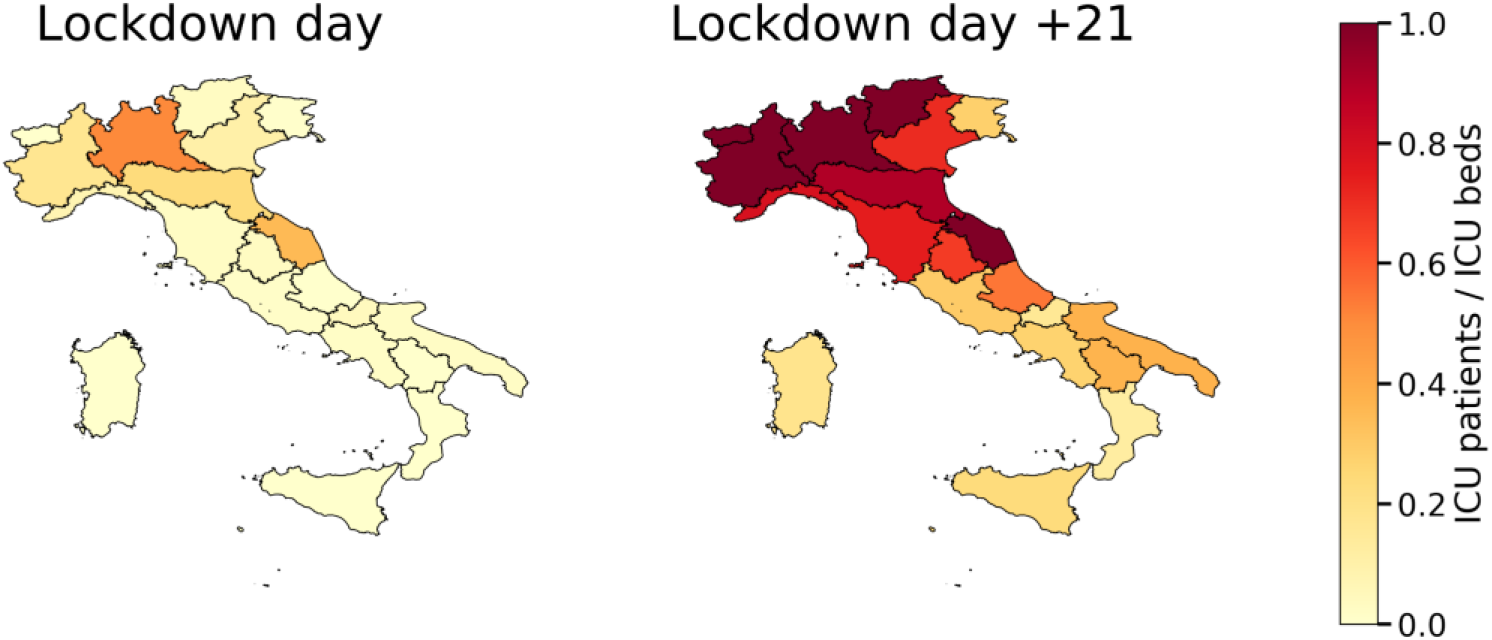
Spatial distribution of COVID-19 ICU patients in Italy. The maps represent the ICUs saturation (number of ICU patients divided by number of ICU beds) at the regional level on March 9^th^ (left panel), when the lockdown was imposed, and twenty-one days later, on March 30^th^ (right panel). Here we use the regional number of ICU beds reported before the onset of the epidemic. The number of ICU beds was increased throughout the epidemic.

### Saturation of ICU can be predicted accurately early on

Here we show that it is possible to predict the date of saturation of the ICUs in a region early on, by using the temporal information about the number of available ICU beds. We focus on Lombardy region. The number of ICU patients in the region grew exponentially for the first ten days, starting from February 24^th^, and then slowed down as it reached the number of available ICU beds. We can predict the ICU saturation date by performing a linear regression of the logarithm of the number of ICU patients, starting from the first four data points (Fig. 4). This result shows that monitoring the ICUs statistics at the beginning of the epidemic allows countries to assess the date of possible saturation of the ICU beds early on. Monitoring the ICUs early in the outbreak is paramount: Lombardy, in Italy, has one the best NHS in the World [11, 12], therefore most countries will face the saturation of their ICU beds at earlier stages of the outbreak. It is worth to note that several factors affect the time to saturate the ICU beds. In particular, the saturation time depends on the connectivity of the population: the more people are connected within a region, the faster the infection diffuses [13]. Therefore, the risk of ICUs saturation is higher for the most developed and connected regions, and Lombardy is the most connected region of Italy. Moreover, if a lockdown measure is imposed, the ICUs saturation time depends on the incubation time of the disease and on the degree of adherence of the population to the lockdown.

### Lockdown benefits for Italy

Here we analyze the effects of the lockdown of the Italian population. Italy imposed two major containment measures: the lockdown on March 9^th^ 2020, and the full lockdown on March 21^st^ 2020, where all non-essential companies and manufacturing plant were closed. These measured helped to avoid the collapse of the whole national health system, yet they could not avoid the saturation of the ICU beds in several Italian regions (Fig. 5).

The effects of the confinement measures become evident with some delay. Specifically, we expect the effect on the number of ICU patients to appear within about two weeks (i.e., the maximum incubation time), and the effect on the number of deaths to appear in about three weeks (i.e., the time from infection to death) [4]. Because of the saturation of the ICU beds in several Italian regions, the number of ICU beds occupied in Italy was lower than the patients that required intensive care. Therefore, we analyze the Italian data excluding the regions where the ICUs had saturated (Fig. 6). This leaves us with 15 of the 20 regions, excluding about 28% of the Italian population. First, we assess the short-term lockdown effects analyzing data up to April 9^th^ (Fig. 6a-b). We find that the growth of ICU patients closely follows a logistic trend, suggesting that the lockdown measures have effectively reduced the spread of the infection (Fig. 6a), as it has been for Hubei region [3]. In the same time interval, the cumulative number of deaths in time can be interpolated well using an exponential curve up to 11 days after the lockdown, and using a line for later data points (Fig. 6b). This implies that the lockdown has spared thousands of lives. Specifically, for the cumulative number of ICU patients, we obtain a good fit using an exponential curve (ICU patients(t) ∝ exp[*r* t], t = days) up to five days after the lockdown (equal to the median incubation time of COVID-19 [14]), and a line (ICU patients(t) ∝ *b* t, t = days) for later data points up to 15 days after the lockdown. Overall, a logistic curve (ICU patients(t) = *k*/(1 + *q* exp[−*p* t]), t = days) represents well the trend of ICU patients in time (Fig. 6a). Finally, we analyze the long-term effects of lockdown, analyzing data up to July 9^th^ (Fig. 6 c-d). The linear growth of deaths continues for at least 45 days after the lockdown declaration (Fig. 6d). In the same period, the ICU patients curve is flattened to zero (Fig. 6c).

**Figure 6.**
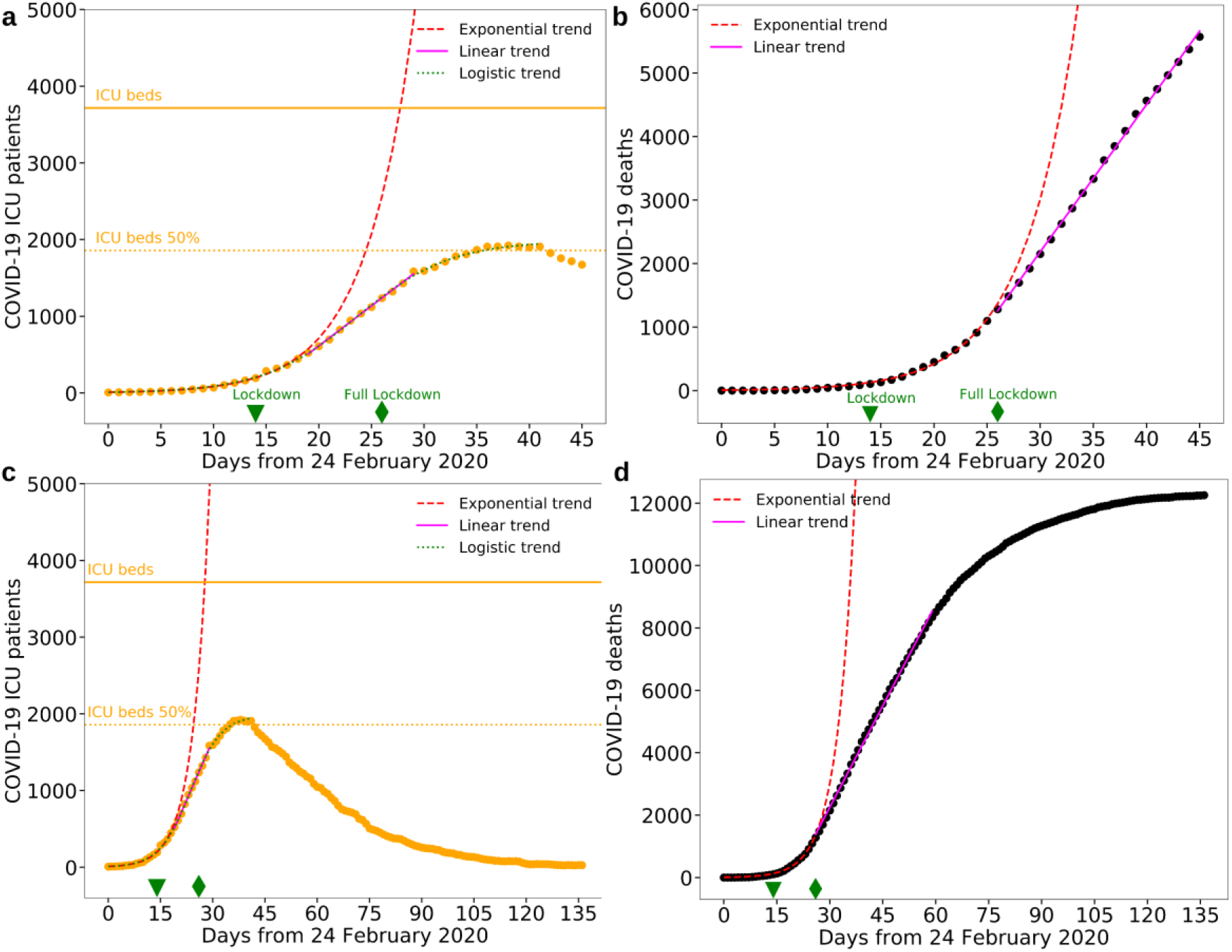
Effect of lockdown in Italy. **a**, The number of ICU patients grows slower than exponential starting from March 14^th^, i.e., five days after the lockdown of March 9^th^ (green triangle). Five days correspond to the current estimate of median incubation time of COVID-19 [14]. We divided the data points into three groups, before March 14^th^, between March 14^th^ and March 24^th^, and up to April 5^th^. We fit the first group with an exponential curve (red dashed line, ICU patients(t) ∝ exp[*r* t], t = days), the second group with a line (magenta line, ICU patients(t) ∝ *b* t, t = days), and the third group with a logistic curve (green dashed line, ICU patients(t) = *k*/(1 + *q* exp[−*p* t]), t = days). The best fit parameters are *r* = 0.213 (95 % CI : 0.197 – 0.229), *b* = 104 (95 % CI : 100 – 108), *k* = 1980 (95 % CI : 1960 – 2000), *q* = 203 (95 % CI : 175 – 231), *p* = 0.225 (95 % CI : 0.219 – 0.231). The root mean square error (RMSE) and normalized-RMSE (n-RMSE) of the exponential fit are 13.80 and 3.1 %, respectively. RMSE and n-RMSE for the linear fit and the logistic fit are 18.73 and 1.8 %, and 20.02 and 1.0 %, respectively. **b**, As for panel **a**, representing the number of deaths. The linear trend starts eleven days after the lockdown. The best fit parameters are *r* = 0.197 (95 % CI : 0.189 – 0.205), and *b* = 230 (95 % CI : 226 – 236). RMSE and n-RMSE for the exponential fit are 20.57 and 1.9 %, respectively; RMSE and n-RMSE for the linear fit are 18.84 and 0.9 %, respectively. **c**-**d**, As for panel **a**-**b**, up to 4 months after the lockdown declaration (July 9^th^). For all panels, we excluded data from the Italian regions where the ICUs saturated, i.e. Lombardy, Piedmont, Marche, Trentino Alto Adige, Valle d’Aosta, which host 28% of the total Italian population. The non-linear least squares problems have been solved using Levenberg-Marquardt algorithm, with NumPy library (Python).

Overall, this analysis suggests that after five days after the lockdown the growth of ICU patients start changing from exponential to linear, and in about 28 days reaches the upper plateau of the logistic trend. Regarding the number of deaths, after about 11 days the growth changes from exponential to linear, and after about 120 days the number approaches a plateau.

## Conclusions

In this work, we analyzed the temporal evolution of COVID-19 outbreak in Italy up to 4 months after the national lockdown declaration of March 9^th^ 2020, and we discussed the short-term (days) and long-term (months) impact of the lockdown in containing the diffusion. The saturation of the ICUs in many Italian regions suggests that containment measures were taken too late. Using Italy as precedent, other countries should impose these confinement measures at earlier stages of the outbreak to be able to protect their population from COVID-19. We show that countries can predict the date of saturation of their ICUs early on, as soon as an exponential growth of intensive care patients is observed, as it was in Lombardy region. The Italian case demonstrates that the national lockdown is effective in reducing the growth of ICU patients. The saturation of ICUs, thus the collapse of a NHS, would be catastrophic, affecting the entire population of a nation. People would die if they needed intensive care for any reason, regardless of their age or their wealth. The saturation of the Italian intensive care units suggests that other governments should act early to contain COVID-19 and consider stronger measures than lockdown, including immediate closing of non-essential companies and manufacturing plants.

Our results can help design better policies to contain the second rapid spread of infections currently observed worldwide. If mild restrictions are not sufficient to contain the infections and the exponential growth of ICU patients, the lockdown remains the only possible measure. We show that at least 5 days are needed from the beginning of lockdown to change the ICU patients growth from exponential to linear, and the increase of ICU patients continues for at least 4 weeks. Moreover, the number of casualties caused by COVID-19 might linearly grow up to 45 days after the lockdown.

We strongly encourage any government to accurately share data, including ICU patients; this data will significantly help the understanding of present and future evolution of the COVID-19 pandemic. Italy has been the first country in Europe stroke by the COVID outbreak, and it has adopted a policy of wide dissemination of open data with detailed spatial structure. Unprecedented measures have been taken by the Italian government, including systematic sharing of ICUs information, and the lockdown of the country. Most likely, the decisions taken in Italy between February 24^th^ and March 21^st^ have saved thousands of lives, not only within national borders.

## Data Availability

The Italian COVID-19 data are available through a GitHub repository managed by the DPC (Dipartimento della Protezione Civile - Presidenza del Consiglio dei Ministri).
For Figure 1, we used World COVID-19 data published by the EU agency for Disease Prevention and Control.
For Figure 1, we also used the dataset maintained by the Center for Systems Science and Engineering (CSSE) at Johns Hopkins University (JHU), for the time series of the Chinese Hubei region.
For Figure 1, we finally used the dataset maintained by the New York Times, for the time series of the US* (New York State + California State).
The number of available beds in ICU in Italy before the onset of the epidemic have been extracted from the dataset published by the Italian Ministry of Health.

https://github.com/pcm-dpc/COVID-19.

https://www.ecdc.europa.eu/en/publications-data/download-todays-data-geographic-distribution-covid-19-cases-worldwide

https://github.com/CSSEGISandData/COVID-19

https://github.com/nytimes/covid-19-data

http://www.dati.salute.gov.it/imgs/C_17_dataset_17_0_upFile.csv

## Authors’ contributions

MS analyzed the data, conceived the manuscript and, together with FL, prepared the figures. All the authors contributed to the interpretation of the data and the discussion of the results. MS and ADC wrote the manuscript, and all the authors reviewed it.

## Acknowledgments

This manuscript has been written with the hope that the heroic resistance showed by the city of Bergamo, the Lombardy region and the whole of Italy will not be needed elsewhere.

The authors want to thank all the Italian nurses, doctors and health-care professionals that are fighting against an invisible enemy as war heroes in dark times. This manuscript is dedicated to the memory of Doctor Li Wenliang, who first tried to warn the World about COVID-19, and all the health-care professionals who died fighting against this virus.

## Availability of data and materials

The Italian COVID-19 data are available through a GitHub repository managed by the DPC (Dipartimento della Protezione Civile - Presidenza del Consiglio dei Ministri): https://github.com/pcm-dpc/COVID-19

For Figure 1, we used World COVID-19 data published by the EU agency for Disease Prevention and Control: https://www.ecdc.europa.eu/en/publications-data/download-todays-data-geographic-distribution-covid-19-cases-worldwide

For Figure 1, we also used the dataset maintained by the Center for Systems Science and Engineering (CSSE) at Johns Hopkins University (JHU), for the time series of the Chinese Hubei region: https://github.com/CSSEGISandData/COVID-19

For Figure 1, we finally used the dataset maintained by the New York Times, for the time series of the US* (New York State + California State): https://github.com/nytimes/covid-19-data

The number of available beds in ICU in Italy before the onset of the epidemic have been extracted from the dataset published by the Italian Ministry of Health: http://www.dati.salute.gov.it/imgs/C_17_dataset_17_0_upFile.csv

